# Early prediction of impending septic shock in children using age-adjusted Sepsis-3 criteria

**DOI:** 10.1101/2020.11.30.20241430

**Authors:** Ran Liu, Joseph L. Greenstein, James C. Fackler, Jules Bergmann, Melania M. Bembea, Raimond L. Winslow

## Abstract

Sepsis is a syndrome which afflicts both adults and children, with many disease courses and diverse outcomes. Understanding of sepsis pathophysiology has changed over time; the Sepsis-3 criteria define sepsis in adults as organ dysfunction, quantified by SOFA score, caused by dysregulated immune response to infection. However, pediatric consensus definitions still utilize the SIRS-based Sepsis-2 criteria, though individual groups have attempted to adapt the Sepsis-3 criteria for children. We evaluate age-adjusted Sepsis-3 criteria on 2,384 pediatric patients admitted to the Johns Hopkins PICU, and apply previously-published methods for early prediction of septic shock. We obtain best early prediction performance of 0.96 AUC, 49.9% overall PPV, and a 5.8-hour median EWT using Sepsis-3 labels based on age-adjusted SOFA score. Through analyses of risk score evolution over time, we corroborate our past finding of an abrupt transition preceding onset of septic shock in children, and are able to stratify pediatric sepsis patients using their first post-threshold-crossing risk score.

## Introduction

Sepsis is a leading cause of in-hospital mortality for both adults and children worldwide^1,2^. Kumar et al. showed that every hour of delayed treatment increased mortality in adult patients by ∼8%^3^. Weiss et al. found this same association in pediatric sepsis patients, and that delayed treatment remains common^4^.

Developing effective guidelines for diagnosis and treatment of sepsis is difficult because sepsis is not a single disease. The Third International Consensus Definitions for Sepsis and Septic Shock (Sepsis-3)^5^ reflect the most recent understanding of sepsis as organ dysfunction caused by dysregulated immune response to infection. However, consensus definitions for pediatric sepsis were last updated in 2005^6^, modeled closely along the Sepsis-2 definitions for adults^7^. Recently, studies attempting to adapt the adult Sepsis-3 criteria for children have been conducted by individual groups^8-10^, and an international expert group published guidelines for the treatment of septic shock and its associated organ dysfunction^11^.

Several computational approaches for early prediction of sepsis and septic shock using electronic health record (EHR) data have been developed with the aim of reducing treatment delays in adult patients^12-14^. We developed a method for predicting an oncoming transition from sepsis to septic shock based on the hypothesis that there exists a physiologically distinct state of sepsis, which we term the “pre-shock state”, and that entry into this state presages the impending onset of septic shock^13^. Through analyses of the temporal evolution of patient state, we previously stratified adult sepsis patients by outcome and interventions received, and discovered that entry into the pre-shock state was marked by a rapid shift in patient physiology occurring within a 30-60 minute timeframe^15^.

In this study, we evaluate age-adapted Sepsis-3 criteria, and apply our previously-published method for early prediction of septic shock on a population of patients admitted to an academic, quaternary center pediatric intensive care unit. We corroborate our past finding of an abrupt transition preceding onset of septic shock in children and are able to stratify sepsis patients using their first post-threshold-crossing risk score.

## Methods

### Study Population

We conducted a retrospective observational cohort study of all patients admitted to the Johns Hopkins Pediatric Intensive Care Unit on or after July 1, 2016, and with discharge prior to January 31, 2018. Patients aged 18 and older were excluded.

### Data Extraction and Processing

Raw data for the Sepsis Quality Improvement Program (SEQUIP) dataset was sourced from an EHR data report and included patient demographics, encounter diagnosis codes, ADT (admit discharge transfer codes with patient room/bed assignments), provider-entered flowsheets (which included nurse-validated vitals and respiratory therapist-validated ventilator settings and measurements), a subset of laboratory results, medication orders, and medication administrations.

The SEQUIP dataset contains data from 2,527 distinct patients admitted to the ICU and 3,125 hospital admissions. Excluding patients aged 18 and older yields our analysis set of 2,384 patients and 2,923 hospital admissions, with an overall sepsis prevalence of 15.88%, as determined by age-adjusted Sepsis-3 criteria^8,9^. Data entries in SEQUIP are comprised of timestamp-value pairs, with ID numbers for patients, hospital admissions, and ICU stays, as well as an ID indicating the feature associated with each value. Complete details of entries corresponding to each physiological variable used in this study are given in the Supplement. 70% of patients were randomly sampled into the training set, and the remaining 30% reserved for testing.

### Labeling Clinical States

Suspected infection was determined using ICD-10 codes. Though Seymour et al.^16^ recommend determining suspected infection from orders of antibiotics and blood cultures, the availability of blood culture data is limited in this dataset, and thus we cannot use this criterion to determine suspected infection. Angus et al. specify ICD-9 codes indicative of suspected infection, used in several prior studies of sepsis^2,12-14^. Mapping between ICD-9 and ICD-10 codes was performed based on the code descriptions (see Supplement, *Infection Criteria*). Comorbidities (Table S2) were computed according to the criteria for the Pediatric Complex Chronic Condition Classification using ICD-10 codes^17,18^.

The Goldstein consensus criteria were evaluated using EHR data from SEQUIP^6^. According to these criteria, sepsis is defined as suspected infection and two or more age-adjusted SIRS criteria (Supplement, *Labeling Clinical States*). Septic shock is defined as sepsis with cardiovascular dysfunction. Labels were re-evaluated each time there was a new observation of clinical data; missing values were imputed using the last observation carried forward, and if no prior observations of a feature were available, the patient was assumed to be within normal ranges for that variable.

According to the Sepsis-3 criteria, sepsis is defined as organ dysfunction consequent to suspected infection. We determine organ dysfunction as a 2-point rise in age-adjusted SOFA score, as defined by Matics et al.^9^ by using the PELOD-2 cutoffs for mean arterial pressure (MAP) and creatinine for cardiovascular and liver SOFA, respectively, and by either a 2-point rise, or a 6-point rise in PELOD-2. Septic shock patients are those with sepsis who were adequately fluid resuscitated, were administered vasopressors, and exhibited a serum lactate concentration greater than 2 mmol/L. Adequate fluid resuscitation was determined using the 2020 SSC pediatric guidelines^11^, and is defined as 40 mL/kg of fluids in the past 3 hours, or having attained the fluid resuscitation target of MAP at the 5^th^ percentile or higher for age, estimated as 1.5 x age in years + 40 mmHg^19^.

### Risk Modeling and Prediction

Risk models were built according to methods previously described, using 26 clinical features extracted from EHR data^13^. MAP and heart rate (HR) were normalized to percentile values by age^19,20^. In order to characterize the pre-shock state, XGBoost and GLM regression models were trained using data from the sepsis clinical state in patients who do not develop septic shock and data from a time window spanning 100 minutes prior to septic shock onset to 1 minute prior to septic shock onset in septic shock patients. Lasso regularization was used for feature selection in GLM models.

For early prediction of septic shock, each patient’s risk score is calculated for each time at which there is EHR data from the beginning of their entries until septic shock onset. Prediction occurs at the first time at which a patient’s risk score exceeds the threshold value. The optimal detection threshold is determined from the training set as the value of the threshold corresponding to the point on the ROC curve closest to the upper left-hand corner. Early warning time (EWT) is defined as the difference from this time until time of septic shock onset. Confidence intervals for performance criteria were estimated using 100 bootstrap iterations.

### Stratification of Sepsis Patients

Stratification of sepsis patients by risk score trajectories was performed as described previously^15^. Risk score trajectories for each patient were computed by applying the XGBoost model at each hour in the time window surrounding time of early prediction using the most recently observed values of each feature. Spectral clustering was applied to these trajectories in order to stratify patients into clusters with similar risk trajectories in the time window following time of early prediction^21^ (Supplement, *Spectral Clustering*).

## Results

### Baseline Statistics

We applied to the SEQUIP dataset four sets of diagnostic criteria for diagnosing sepsis and septic shock. These criteria were the 2005 Goldstein consensus criteria and three others evaluated by Schlapbach et al.^8^. Using these labels, we computed the prevalence and mortality rate in each cohort (Table 2). Depending on the criteria used, the prevalence of sepsis varies between 5.5% to 35%, and the prevalence of septic shock ranges from 2% to almost 20%. We find that sepsis patients labeled using age-adjusted Sepsis-3 criteria have greater mortality than those labeled using the Goldstein criteria. When using age-adjusted SOFA scores to determine clinical state labels, mortality in both sepsis cohorts (sepsis without shock and septic shock patients) is higher than in the corresponding cohort determined using the SIRS-based Goldstein criteria. When organ dysfunction is defined by an increase of 6 points or greater in PELOD-2 score, this results in the lowest prevalence of sepsis, and the highest mortality in both sepsis cohorts. The Goldstein criteria result in the highest prevalence of sepsis, but the lowest mortality in both sepsis cohorts.

**Table 1:** Baseline statistics of SEQUIP dataset. Sepsis cohorts are determined using age-adjusted SOFA score^8^.

| **Most severe clinical state reached** | **No sepsis** | **Sepsis without septic shock** | **Sepsis leading to septic shock** | **Overall** |
| --- | --- | --- | --- | --- |
| Admissions | 2459 (84.13%) | 353 (12.08%) | 111 (3.80%) | 2923 |
| Patients | 1984 (83.22%) | 294 (12.33%) | 106 (4.45%) | 2384 |
| PICU Stays | 2944 (84.4%) | 407 (11.68%) | 135 (3.87%) | 3486 |
| In-hospital mortality | 35 (1.42%) | 11 (3.12%) | 18 (16.22%) | 64 (2.19%) |
| Gender | 1166 M (58.77%),<br>818 F (41.23%) | 182 M (61.9%),<br>112 F (38.1%) | 56 M (52.83%),<br>50 F (47.17%) | 1404 M (58.89%),<br>980 F (41.11%) |
| Median ICU stay length, days (IQR) | 1.53 (0.88, 2.90) | 3.79 (1.70, 8.16) | 12.25 (5.67, 26.91) | 1.73 (0.92, 3.73) |
| Median age, years (IQR) | 5.33 (1.27, 11.68) | 2.08 (0.78, 5.44) | 1.20 (0.01, 9.03) | 4.55 (1.08, 11.14) |

**Table 2:** Comparison of diagnostic criteria: hospital admission counts and proportion of all admissions represented by each cohort in parentheses **(A)**, and mortality in each cohort **(B)**.

| **Criteria** | **Non-sepsis** | **Sepsis without shock** | **Septic shock** |
| --- | --- | --- | --- |
| Goldstein | 1831 (62.58%) | 488 (16.68%) | 607 (20.75%) |
| Age-adjusted SOFA | 2459 (84.13%) | 353 (12.08%) | 111 (3.80%) |
| PELOD-2 (2 points) | 1912 (65.35%) | 878 (30.01%) | 136 (4.65%) |
| PELOD-2 (6 points) | 2767 (94.57%) | 101 (3.45%) | 58 (1.98%) |

| **Criteria** | **Non-sepsis** | **Sepsis without shock** | **Septic shock** |
| --- | --- | --- | --- |
| Goldstein | 33 (1.80%) | 0 (0.00%) | 31 (5.11%) |
| Age-adjusted SOFA | 35 (1.42%) | 11 (3.12%) | 18 (16.22%) |
| PELOD-2 (2 points) | 33 (1.73%) | 13 (1.48%) | 18 (13.24%) |
| PELOD-2 (6 points) | 42 (1.52%) | 10 (9.90%) | 12 (20.69%) |

### Early Prediction of Septic Shock

Figure 1 shows risk score trajectories using XGBoost^22^ from a patient who does (Figure 1A) and a patient who does not (Figure 1B) develop septic shock.

**Figure 1:**
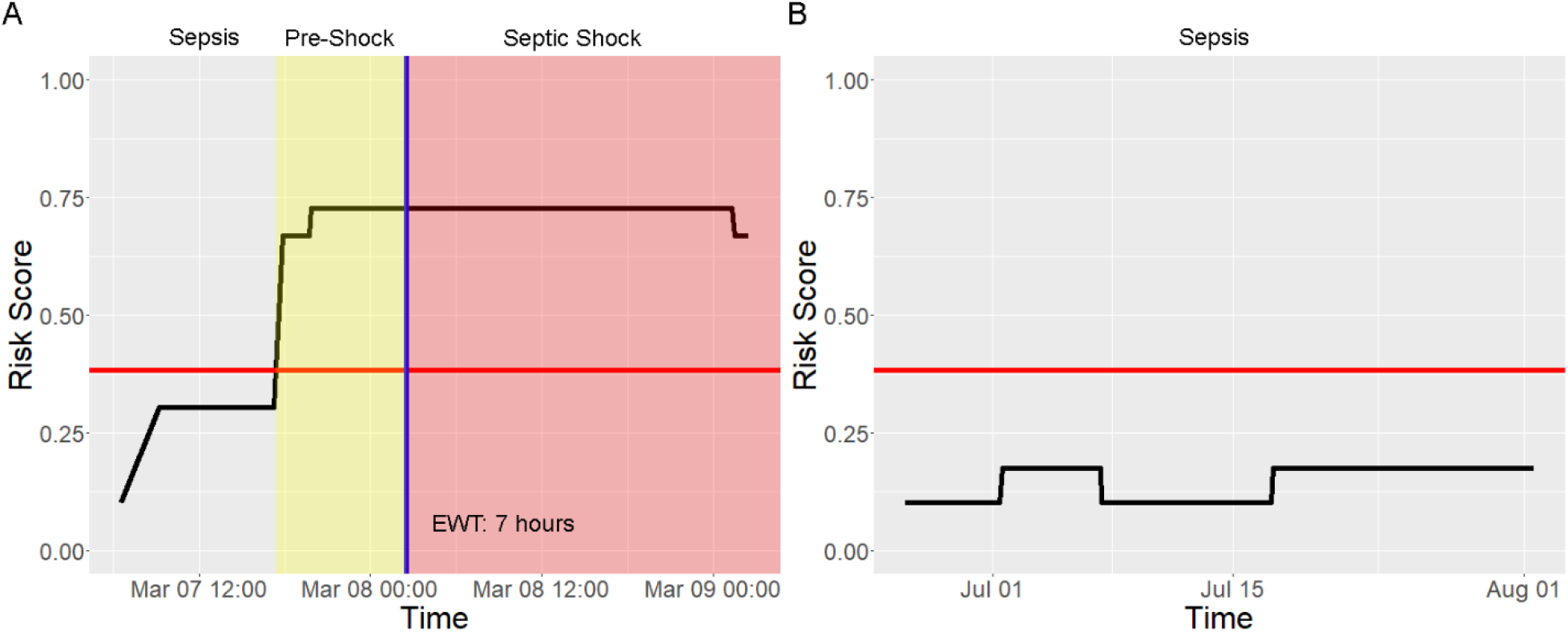
Example risk trajectories for (A) a patient who developed septic shock and (B) a non-shock sepsis patient. Threshold for early prediction is indicated by the red horizontal line, and time of septic shock onset is indicated by the blue vertical line.

The performance of our method for early prediction of septic shock was evaluated on the SEQUIP dataset using two machine learning methods, GLM^23^ and XGBoost^22^. Figure 2 shows the receiver operating characteristic (ROC) curves and performance metrics, for clinical labels produced using the pediatric Sepsis-3 criteria with age-adjusted SOFA scores. We evaluated performance in early prediction using all sets of clinical criteria, and found that best performance overall was obtained with labels using age-adjusted SOFA scores (Figure S2). Of the two machine learning methods, XGBoost yields greater performance, with 0.96 AUC and 49.9% average PPV, with a shorter median EWT of 5.8 hours compared to 0.92 AUC, 41.0% PPV, and 10.5 hours median EWT when using GLM.

**Figure 2:**
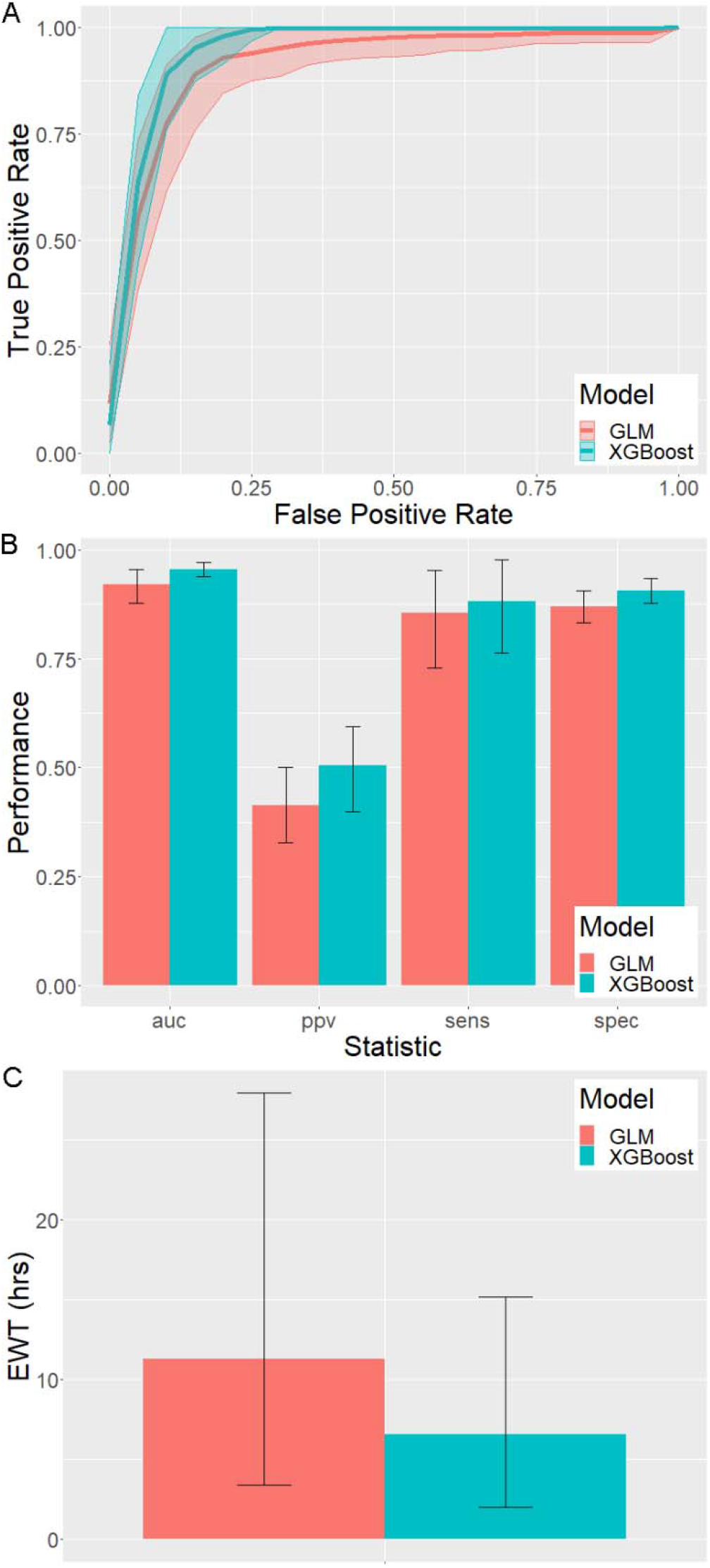
Performance of early prediction of septic shock in pediatric patients: (A) ROC curves and 90% confidence intervals, (B) area under the ROC curve (AUC), positive predictive value (PPV), sensitivity, and specificity for GLM and XGBoost where error bars indicate 90% confidence intervals, and (C) early warning time (EWT) for GLM and XGBoost where error bars indicate 90% confidence intervals.

Both XGBoost and GLM models are interpretable by examining the importance of each feature in the models. Feature importance is given in Table S8. XGBoost is a decision tree gradient boosting method. For XGBoost, gain is the average relative increase in performance after the addition of a feature to a model. Coverage is the relative frequency at which decision tree nodes which split on a particular feature are reached. Frequency is the relative frequency at which a feature appears in decision trees. For GLM, exponentiated coefficients directly explain the magnitude of the contribution of each feature to the risk score and can be interpreted as odds ratios. For example, the exponentiated coefficient of lactate is 2.14. Therefore, a patient with serum lactate concentration 1 standard deviation above the population mean is approximately twice as likely to develop septic shock than a patient with average serum lactate.

### Stratification of Sepsis Patients

As previously published^13^, our approach to early prediction allows for the calculation of a patient-specific PPV. This is performed based on the first value of risk score that exceeds the threshold. These values were binned into quartiles, and PPV was computed in each bin, yielding an estimate of the probability that a prediction of impending septic shock onset for a patient whose risk score falls into the specified range is a true positive (Table S9). In higher quartiles, the likelihood that predictions are true positives is greater than in lower quartiles, with PPV as large as 80%.

We repeated our analyses of risk score trajectories for stratification of sepsis patients^15^ on the SEQUIP dataset. Spectral clustering^21^ of risk score trajectories in the window surrounding early prediction yielded two clusters (Figure 3). Patient risk trajectories are largely indistinguishable prior to the time of early prediction. Risk scores increases abruptly at the time of threshold crossing for all patients, and then clusters diverge subsequent to that. Clusters stratify by the prevalence of septic shock, time to septic shock onset, and the proportion of patients who are adequately fluid resuscitated prior to time of early warning (Table S10). Differences in mortality between these clusters, however, are not statistically significant.

**Figure 3:**
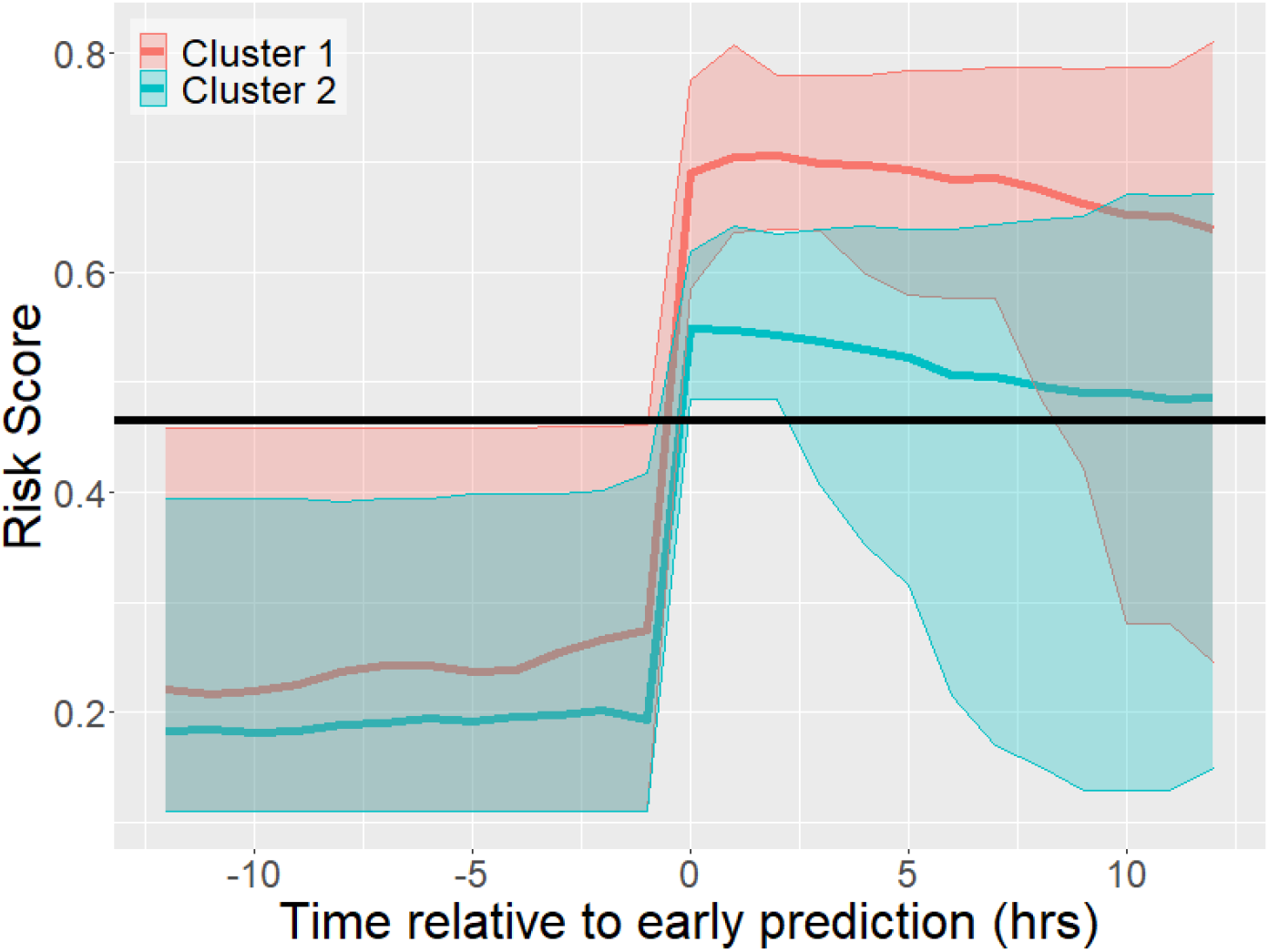
Clusters of risk score trajectories in the window surrounding time of early warning.

## Discussion

### Pediatric Sepsis Criteria

The shortcomings of SIRS-based criteria for sepsis are well-known. The presence of SIRS is not specific for infection, and in adults, over 90% of intensive care patients meet the criteria for SIRS^24-26^. The Sepsis-3 criteria for adults redefined sepsis as infection resulting in organ dysfunction, determined by an increase of at least 2 points in SOFA score^27,28^. Consequently, there has been strong interest in redefining pediatric sepsis on the basis of organ dysfunction. Shortly after the publication of the Sepsis-3 criteria, Leclerc et al. suggested the use of PELOD-2 scores in children with suspected infection^10^, and Matics et al. suggested the use of an age-adjusted SOFA score. Our findings support the usage of pediatric Sepsis-3 criteria based on an age-adjusted SOFA score, as these criteria achieve the greatest performance in early prediction, with an AUC of 0.96 and overall PPV of 49.9%. Furthermore, we corroborate the findings of other groups that age-adjusted SOFA and PELOD-2 have greater validity in stratifying patients by metrics of disease severity than the SIRS-based Goldstein criteria (Figure S4)^8^.

### Prediction Method

Just as in adults, the goal of early prediction of septic shock in pediatric patients is to provide clinicians with a time window of intervention for patients at high risk of deteriorating from sepsis to septic shock, enabling more timely treatment and possibly preventing development of shock. The premise of our method for early prediction of septic shock is the existence of a pre-shock state. In this study, we validate our previously published methodology by demonstrating the role of this pre-shock state in early prediction. We achieve high performance of 0.96 AUC, 49.9% overall PPV, and a 5.8-hour median early warning time. Model performance using XGBoost^22^ is higher than that in our prior study on data from adult patients, and is higher than that obtained using GLM^23^. XGBoost uses gradient boosting of decision trees, and thus is able to learn nonlinear associations between features and risk. This is likely the source of its improved performance compared to GLM, which is consistent with our previous results.

In our previous work, we introduced the notion of patient-specific positive predictive value, where patients were stratified into 10 bins based on their first post-threshold crossing value of risk^13^. Due to the much smaller number of patients in the SEQUIP dataset compared to the MIMIC-III and eICU datasets (2,650 vs 38,418 vs 139,367), and the lower prevalence of sepsis and septic shock in the pediatric population, we cannot achieve the same level of resolution in estimating patient-specific PPV, as there would be too few patients to estimate the PPV in each stratum. Therefore, we stratified positive predictions into quartiles. However, we demonstrate that our method for estimating the confidence of a positive prediction remains applicable in pediatric sepsis patients. Being able to estimate the confidence level of a prediction enables clinicians to act only on higher-confidence predictions, or take different courses of action informed with the likelihood that a patient develops septic shock.

### Stratification of Patients

Previously, using spectral clustering of patient risk score trajectories, we further elucidated the pre-shock state, finding that entry into the state was marked by a rapid transition from low to high risk. Prior to entry, sepsis patient physiology was indistinguishable between the low and high risk clusters of patients^15^. However, after the occurrence of this event, patient risk trajectories diverged, and stratified patients by risk of septic shock, mortality, time to septic shock onset, and treatments received. We did not find the difference in mortality between clusters to be statistically significant in pediatric patients; this is likely due to the relatively small size of our data set. The limited number of patients in our dataset also causes further difficulties in applying clustering to risk trajectories. If we cluster patients based on risk trajectories in the 12 hours following early prediction, then the space over which we are clustering is 13-dimensional; such a space is sparsely populated by the roughly 200 trajectories which exceed the prediction threshold ^21^. Nonetheless, this does not invalidate our finding that a very rapid transition in risk score at the time of threshold crossing occurs in pediatric as well as adult sepsis patients, and that risk score trajectories are able to stratify pediatric sepsis patients by risk of septic shock, time to shock onset, and treatments received. These findings support our postulation of the pre-shock state, and that the entry into septic shock is extremely rapid in both adult and pediatric sepsis patients. The rapid nature of the transition further indicates the necessity of automated methods for the detection and prediction of septic shock.

### Model Interpretation

It is possible to determine the relative importance of features in both XGBoost and GLM models. Both models share 4 out of their top 5 features in common: lactate, respiratory SOFA, PaO_2_, and BUN. Lactate is the most important feature in both models, as was true in our previous study of adult sepsis patients. These findings align with existing literature on the pathophysiology of sepsis and septic shock. Elevated serum lactate indicates reduced tissue perfusion, and predicts mortality in patients with infections^29,30^. Increased respiratory SOFA and abnormal PaO_2_ are both associated with respiratory dysfunction^27^. BUN levels are associated with kidney function, which is known to be affected in pediatric sepsis^31^.

### Limitations

We algorithmically determine sepsis and septic shock labels according to the Sepsis-3 consensus definitions, using age-adjusted SOFA score or PELOD-2 in order to determine organ dysfunction, and the Surviving Sepsis Campaign^32^ guidelines to determine when patients are adequately fluid resuscitated. Therefore, limitations inherent to these labels are also limitations of the study. For example, the Infectious Disease Society of America points out that determining septic shock in adults using adequate fluid resuscitation as a criterion results in some ambiguity and disagreement in interpretation as to the exact time of onset^33^. Deutschman further remarks that the Sepsis-3 criteria may not precisely reflect the pathophysiology of sepsis, which may become life-threatening through mechanisms other than organ dysfunction^34^. These limitations would also apply in our analysis of age-adjusted Sepsis-3 in pediatric sepsis patients. Completeness of data records in the EHR may also influence the accuracy of our labels (e.g. availability of culture and antibiotic data to determine suspected infection), as well as the accuracy of our predictions. Furthermore, our dataset is limited to a relatively small set of patients from a single center of care. While we corroborate results obtained by other research groups in other cohorts of patients, greater validity could be achieved via a multi-center analysis encompassing a greater number of patients treated within diverse settings.

## Data Availability

Data referred to in this manuscript are not publicly available.

