## Supplementary Materials for "Early prediction of impending septic shock in children using age-adjusted Sepsis-3 criteria"

**Materials and Methods:**

*Infection Criteria*

Angus et al. specify ICD-9 codes indicative of suspected infection (*1*), whereas diagnoses in SEQUIP are provided in the form of ICD-10 codes. Mapping between ICD-10 codes and ICD-9 codes was done on the basis of ICD-10 code descriptions. ICD-10 codes with descriptions containing text matching categories specified in Angus et al. were considered indicative of suspected infection (*1*). A full list of descriptions is given in Table S5.

*Labeling Clinical States*

We applied four sets of criteria for determining sepsis and septic shock in pediatric patients: the Sepsis-2-based Goldstein criteria (*2*), and three sets of Sepsis-3 criteria adapted for sepsis patients. The Goldstein criteria define sepsis as suspected infection, as well as the fulfillment of 2 or more SIRS criteria (*2*). A full list of age-adjusted criteria for variables used in determining SIRS is given in Table S6. For SIRS, septic shock is defined as sepsis as well as cardiovascular dysfunction. Cardiovascular dysfunction is defined as hypotension, which is determined as a mean arterial pressure (MAP) below the 5^th^ percentile for age, computed as 1.5 × (age in years) + 40 mmHg (*3*), or the administration of vasopressors, or a lactate concentration of 2.5 mmol/L and urine output <0.5 mL/kg/hr (*2*).

Sepsis-3 (*4*) defines sepsis as organ dysfunction consequent to suspected infection. Our three sets of Sepsis-3 criteria are defined by the different methods by which we determine the presence of organ dysfunction. The first is using age-adjusted SOFA score (*5, 6*), where organ dysfunction is determined by a 2-point increase in age-adjusted SOFA score. Age-adjusted SOFA score, as suggested by Matics et al., uses the PELOD-2 age cutoffs for MAP and creatinine (*6*). A full list of age cutoffs for variables used in calculating age-adjusted SOFA score is given in Table S7. The second and third criteria determine organ dysfunction using PELOD-2 score, where either a 2-point increase or a 6-point increase in PELOD-2 score indicates organ dysfunction (*7, 8*). Septic shock patients are those that have sepsis, have received adequate fluid resuscitation, require vasopressors to maintain a mean arterial blood pressure of at least 65 mmHg, and have a serum lactate concentration >2 mmol/L. Adequate fluid resuscitation was determined using the 2016 SSC guidelines (*9*): adequate fluid resuscitation is defined as having received 30 mL/kg of fluids, or having attained fluid resuscitation targets of 0.5 mL/kg/hr urine output, or MAP of at least 65 mmHg.

**Discussion:**

*Stratification of Patients*

One method for determining the optimal number of clusters when performing spectral clustering is to use the eigengap heuristic. This procedure selects k such that the gap between the k-th and (k+1)-th eigenvalues of the graph Laplacian is relatively large compared with gaps between all other consecutive pairs of eigenvalues. Geometrically, by the Davis-Kahan theorem, this guarantees that the eigenvectors of the graph Laplacian are robust to small perturbations in the data (*10*). Intuitively, this means that the results of spectral clustering for a selected value of k will be robust to small changes in the data, which is one common measure of goodness of fit for clustering algorithms (*11*).

However, in the case of our data, this method does not yield a clear optimal number of clusters as there are gaps between several eigenvalues of similar size (Figure S1). This is likely due to the small size of our dataset. Thus, we showed results for 2 clusters, as the minimum number necessary to demonstrate the existence of a relationship between risk score trajectories and outcome.

**Supplementary Figures:**

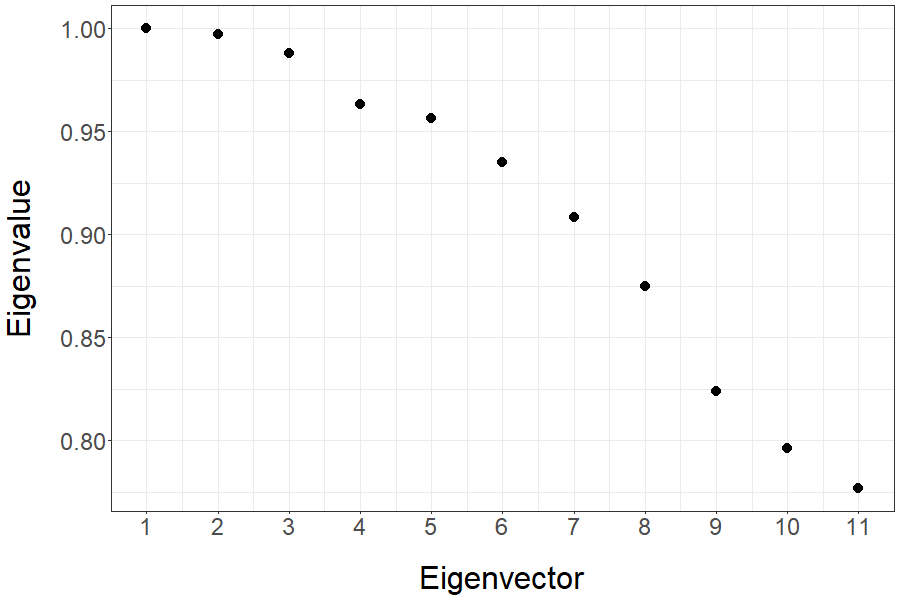

**Figure S1:** Eigenvalues of Graph Laplacian of risk trajectories following time of early prediction.

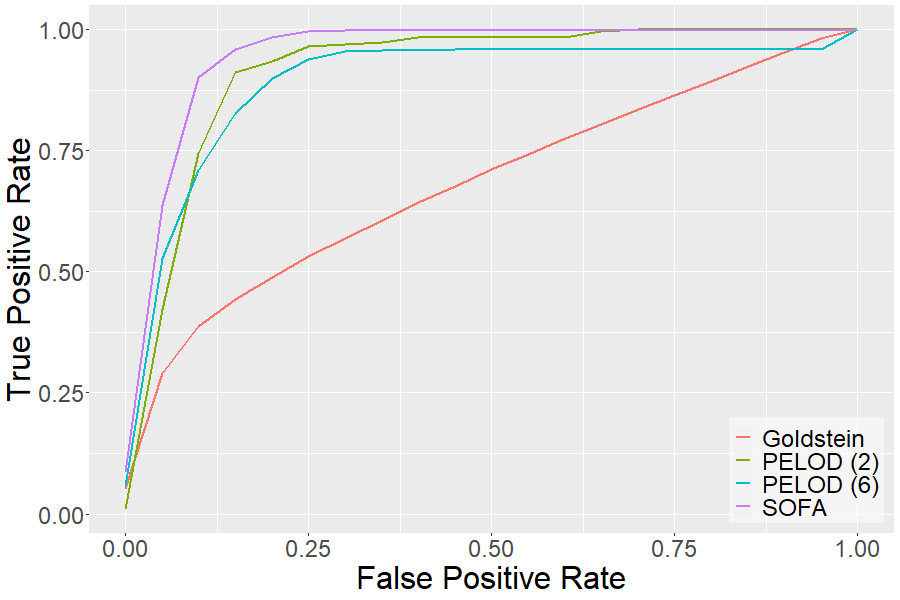

**Figure S2:** Average ROC curves for early prediction using different clinical criteria for labeling of sepsis and septic shock patients.

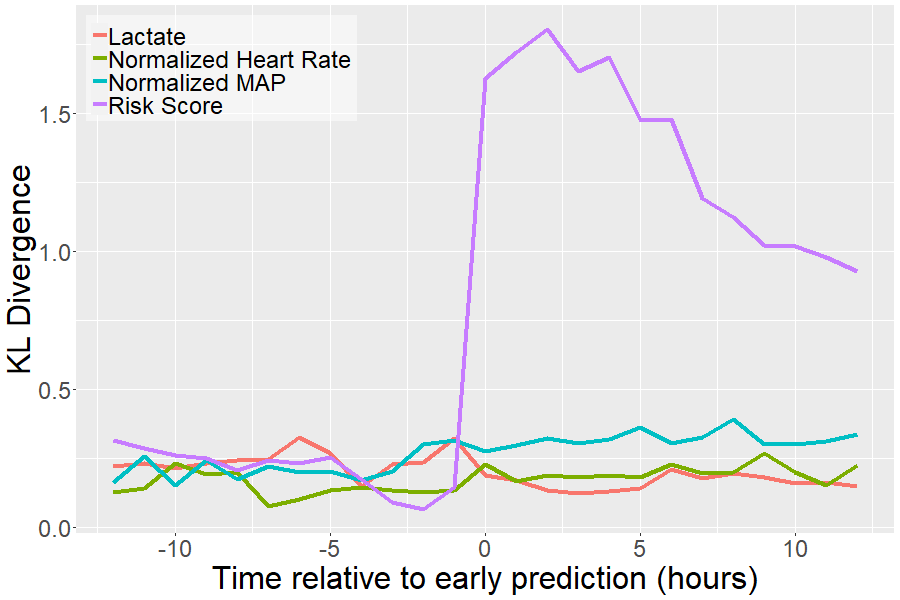

**Figure S3:** Kullback-Leibler divergences of risk score, heart rate, lactate, and MAP in SEQUIP.

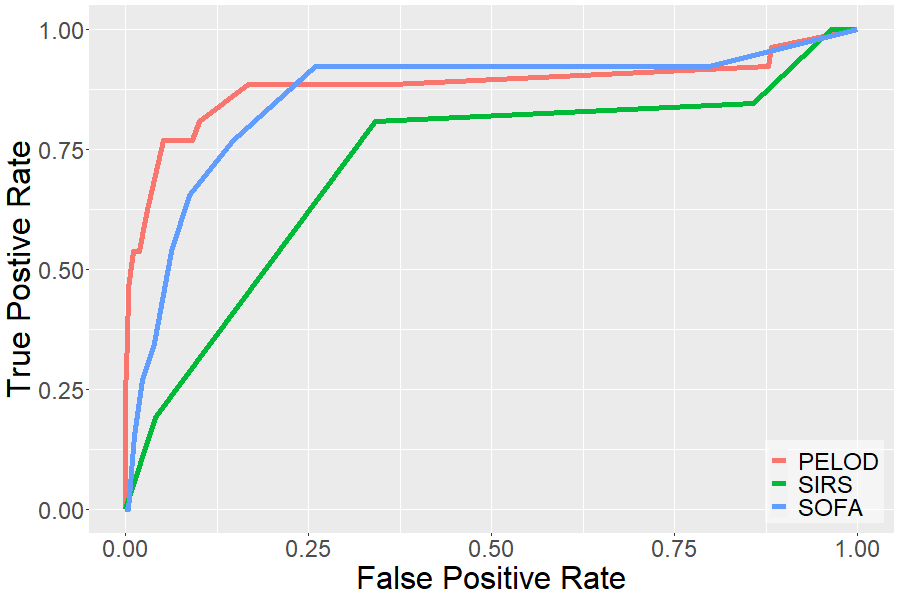

**Figure S4:** ROC curves for prediction of mortality using most severe value of PELOD-2, age-adjusted SOFA, and SIRS.

**Supplementary Tables:**

**Table S1:** Queried Items in SEQUIP

| **Feature** | **Table** | **Items** |
| --- | --- | --- |
| Heart Rate | Flowsheet | 8 |
| Mean Arterial Pressure | Flowsheet | 301250,301360 |
| Respiratory Rate | Flowsheet | 9 |
| Temperature | Flowsheet | 6 |
| CVP | Flowsheet | 301370 |
| PaO_2_ | Labs | 2000000122 |
| FiO2 | Flowsheet | 301550,1601063025,3040019917,304064870 |
| GCS | Flowsheet | 30440104971,30440104966 |
| Bilirubin | Labs | 2000000107 |
| Platelets | Labs | 2000000008 |
| Creatinine | Labs | 2000000751 |
| Lactate | Labs | 2000000900 |
| BUN | Labs | 2000000100 |
| pH | Labs | 2000000120 |
| WBC | Labs | 2000000722 |
| PaCO_2_ | Labs | 2000000121 |
| Hemoglobin | Labs | 2000000897 |
| Hematocrit | Labs | 2000000003 |
| Potassium | Labs | 2000000096 |

**Table S2:** Pediatric Complex Chronic Conditions in SEQUIP

| **Comorbidity** | **Non-sepsis** | **Sepsis** | **Shock** | **Overall** |
| --- | --- | --- | --- | --- |
| Neurological/neuromuscular | 21.7% | 36.0% | 37.0% | 24.1% |
| Cardiovascular | 22.9% | 40.0% | 66.4% | 26.9% |
| Respiratory | 7.8% | 20.8% | 26.9% | 10.2% |
| Renal and urologic | 8.3% | 13.9% | 28.6% | 9.9% |
| Gastrointestinal | 16.2% | 37.9% | 47.1% | 20.2% |
| Hematological/immunological | 4.5% | 15.8% | 16.8% | 6.4% |
| Metabolic | 12.7% | 31.2% | 49.6% | 16.6% |
| Congenital/genetic | 9.9% | 21.9% | 26.1% | 11.8% |
| Malignancy | 5.5% | 16.7% | 13.4% | 7.2% |
| Premature and neonatal | 4.2% | 13.9% | 33.6% | 6.6% |
| Technology dependence | 25.0% | 48.3% | 62.2% | 29.5% |
| Transplant | 1.12% | 3.2% | 5.9% | 1.6% |

**Table S3:** Availability of EHR data in SEQUIP

| **Feature** | **% of Patients with at least 1 entry** | **Average time (hrs) between observations (mean/median)** |
| --- | --- | --- |
| Heart Rate | 99.90 | 1.13/1.00 |
| MAP | 99.58 | 1.43/1.00 |
| Respiratory Rate | 99.90 | 1.20/1.00 |
| Temperature | 99.87 | 2.02/2.00 |
| CVP | 13.60 | 0.81/0.45 |
| PaO_2_ | 26.34 | 7.64/3.20 |
| FiO_2_ | 71.33 | 1.41/1.00 |
| GCS | 47.58 | 2.48/2.00 |
| Bilirubin | 51.42 | 39.12/23.72 |
| Platelets | 68.51 | 28.15/20.40 |
| Creatinine | 0.22 | 312.42/312.42 |
| Lactate | 37.18 | 11.62/3.95 |
| BUN | 75.10 | 25.78/21.63 |
| Arterial pH | 25.92 | 7.03/3.48 |
| WBC | 29.82 | 75.88/12.41 |
| PaCO_2_ | 26.34 | 7.46/3.17 |
| Hemoglobin | 32.93 | 14.39/3.65 |
| Hematocrit | 69.18 | 28.49/20.90 |
| Potassium | 73.41 | 26.01/21.68 |

**Table S4:** Central Tendency Measures of Patient Physiological Data in SEQUIP

| **Feature** | **Mean** | **Median** |
| --- | --- | --- |
| Weight (kg) | 27.66 | 18.8 |
| Heart Rate (bpm) | 122.45 | 124.0 |
| MAP (mmHg) | 68.87 | 68.0 |
| Respiratory Rate (bpm) | 33.45 | 31.0 |
| Temperature (°F) | 98.17 | 98.2 |
| CVP (mmHg) | 11.21 | 10.0 |
| PaO_2_ (mmHg) | 130.13 | 107.0 |
| FiO_2_ (%) | 56.46 | 50.0 |
| GCS | 11.88 | 14.0 |
| Bilirubin (mg/dL) | 1.14 | 0.50 |
| Platelets (1000/µL) | 209.28 | 162.0 |
| Creatinine (mg/dL) | 0.88 | 0.75 |
| Lactate (mmol/L) | 2.05 | 1.50 |
| BUN (mg/dL) | 16.56 | 11.0 |
| Arterial pH | 7.37 | 7.38 |
| WBC (1000/µL) | 10.13 | 5.0 |
| PaCO_2_ (mmHg) | 46.34 | 44.0 |
| Hemoglobin (g/dL) | 11.19 | 11.0 |
| Hematocrit (%) | 30.71 | 30.0 |
| Potassium (mmol/L) | 4.09 | 4.0 |

**Table S5:** ICD-10 codes mapping to categories specified as indicative of suspected infection.

| **Description** |  |
| --- | --- |
| cholera | leptospirosis |
| typhoid | vincent's angina |
| paratyphoid fever | yaws |
| salmonella | pintadermatophytosis |
| shigellosis | candidiasis |
| food poisoning | coccidiodomycosis |
| infection | histoplasmosis |
| tuberculosis | mycoses |
| plague | meningitis |
| tularemia | phlebitis |
| anthrax | pericarditis |
| brucellosis | endocarditis |
| glanders | thrombophlebitis |
| meloidosis | sinusitis |
| rat-bite fever | pharyngitis |
| bacterial zoonoses | tonsilitis |
| leprosy | laryngitis |
| mycobacteria | tracheitis |
| diphtheria | pneumonia |
| whooping cough | bronchitis |
| streptococcal throat | bronchiectasis |
| scarlet fever | empyema |
| erysipelas | diverticulitis |
| tetanus | abscess |
| septicemia | cellulitis |
| sepsis | lymphadenitis |
| septic shock | osteomyelitis |
| bacteria | bactaremia |
| syphilis |  |

**Table S6:** Age-adjusted SIRS criteria.

| **Age Range** | **Temperature (°C)** | **Heart Rate (beats/min)** | **Respiratory Rate (breaths/min)** | **WBC Count (k/mm^3^)** |
| --- | --- | --- | --- | --- |
| 0-1 week | <36 or >38.5 | >180 or <100 | >50 | >34 |
| 1 week-1 month | <36 or >38.5 | >180 or <100 | >40 | >19.5 or <5 |
| 1 month-1 year | <36 or >38.5 | >180 or <90 | >34 | >17.5 or <5 |
| 2-5 years | <36 or >38.5 | >140 | >22 | >15.5 or <6 |
| 6-12 years | <36 or >38.5 | >130 | >18 | >13.5 or <4.5 |
| 13-17 years | <36 or >38.5 | >110 | >14 | >11 or <4.5 |

**Table S7:** Age-adjusted SOFA Score

A. Mean Arterial Pressure Age Cutoffs for Cardiovascular SOFA Score

| Age \ Cardiovascular SOFA Score | 0 | 1 |
| --- | --- | --- |
| <1 month | MAP <46 mmHg | MAP ≥46 mmHg |
| 1 - <12 months | MAP <55 mmHg | MAP ≥55 mmHg |
| 12 - <24 months | MAP <60 mmHg | MAP ≥60 mmHg |
| 24 - <60 months | MAP <62 mmHg | MAP ≥62 mmHg |
| 60 - <144 months | MAP <65 mmHg | MAP ≥65 mmHg |
| ≥144 months | MAP <67 mmHg | MAP ≥67 mmHg |

B. Creatinine Age Cutoffs for Kidney SOFA Score

| Age \ Kidney SOFA Score | 0 | 2 |
| --- | --- | --- |
| <1 month | Creatinine ≤69 µmol/L | Creatinine >69 µmol/L |
| 1 - <12 months | Creatinine ≤22 µmol/L | Creatinine >22 µmol/L |
| 12 - <24 months | Creatinine ≤34 µmol/L | Creatinine >34 µmol/L |
| 24 - <60 months | Creatinine ≤50 µmol/L | Creatinine >50 µmol/L |
| 60 - <144 months | Creatinine ≤58 µmol/L | Creatinine >58 µmol/L |
| ≥144 months | Creatinine ≤92 µmol/L | Creatinine >92 µmol/L |

**A. XGBoost**

| **Feature** | **Gain** | **Cover** | **Frequency** |
| --- | --- | --- | --- |
| Lactate | 0.21 | 0.17 | 0.07 |
| Respiratory SOFA | 0.08 | 0.12 | 0.02 |
| Heart Rate | 0.08 | 0.01 | 0.07 |
| PaO_2_ | 0.06 | 0.06 | 0.05 |
| BUN | 0.06 | 0.05 | 0.06 |
| Bilirubin | 0.06 | 0.02 | 0.05 |
| MAP | 0.05 | 0.12 | 0.07 |
| Urine Output | 0.05 | 0.06 | 0.06 |
| Hematocrit | 0.04 | 0.06 | 0.06 |
| Temperature | 0.04 | 0.11 | 0.06 |

**B. GLM**

| **Feature** | **Coefficient** | **SE** | **Odds Ratio** |
| --- | --- | --- | --- |
| Lactate | 0.76 | 0.01 | 2.14 |
| Respiratory SOFA | 0.73 | 0.01 | 2.08 |
| PaO_2_ | 0.48 | 0.02 | 1.62 |
| BUN | 0.44 | 0.01 | 1.55 |
| MAP | -0.36 | 0.02 | 0.70 |
| WBC | 0.35 | 0.02 | 1.42 |
| Heart Rate | 0.31 | 0.01 | 1.36 |
| Urine Output | 0.29 | 0.02 | 1.34 |
| FiO_2_ | -0.26 | 0.01 | 0.77 |

**Table S8:** Feature importance using (A) XGBoost and (B) GLM for top 10 selected features, sorted in descending order of absolute importance.

| **Percentiles** | **False Positives (1-PPV)** | **True Positives (PPV)** |
| --- | --- | --- |
| **0-25** | 12 (66.7%) | 6 (33.3%) |
| **26-50** | 12 (70.6%) | 5 (29.4%) |
| **51-75** | 11 (57.9%) | 8 (42.1%) |
| **76-100** | 3 (20.0%) | 12 (80.0%) |

**Table S9:** Stratification of patients by first post-threshold crossing value of risk score.

| **Cluster** | **Size** | **Shock Prevalence** | **Mortality** | **Median EWT** | **% Patients Adequately Fluid Resuscitated** | **% Patients Treated with Vasopressors** |
| --- | --- | --- | --- | --- | --- | --- |
| 1 | 108 | 65.7% | 11.1% | 3.0 hours | 69.8% | 46.2% |
| 2 | 120 | 31.7% | 10.8% | 10.4 hours | 80.7% | 47.2% |

**Table S10:** Outcome characteristics of clusters in Figure 3.
